# Distinct psychopathology profiles in patients with epileptic seizures compared to non-epileptic psychogenic seizures

**DOI:** 10.1101/19003038

**Authors:** Albert D Wang, Michelle Leong, Benjamin Johnstone, Genevieve Rayner, Tomas Kalincik, Izanne Roos, Patrick Kwan, Terence J O’Brien, Dennis Velakoulis, Charles B Malpas

## Abstract

**Objective:** Similarities in clinical presentations between epileptic seizures (ES) and psychogenic non-epileptic seizures (PNES) produces a risk of misdiagnosis. Video-EEG monitoring (VEM) is the diagnostic gold standard, but involves significant cost and time commitment, suggesting a need for efficient screening tools.

**Methods:** 628 patients were recruited from an inpatient VEM unit; 293 patients with ES, 158 with PNES, 31 both ES and PNES, and 146 non-diagnostic. Patients completed the SCL-90-R, a standardised 90-item psychopathology instrument. Bayesian linear models were computed to investigate whether SCL-90-R domain scores or the overall psychopathology factor *p* differed between groups. Receiver operating characteristic (ROC) curves were computed to investigate the PNES classification accuracy of each domain score and *p*. A machine learning algorithm was also used to determine which subset of SCL-90-R items produced the greatest classification accuracy.

**Results:** Evidence was found for elevated scores in PNES compared to ES groups in the symptom domains of anxiety (*b* = 0.47, 95%HDI = [0.10, 0.80]), phobic anxiety (*b* = 1.32, 95%HDI = [0.98, 1.69]), somatisation (*b* = 0.84, 95%HDI = [0.49, 1.20]), and the general psychopathology factor *p* (*b* = 1.35, 95%HDI = [0.86, 1.82]). Of the SCL-90-R domain scores, somatisation produced the highest classification accuracy (AUC = 0.74, 95%CI = [0.69, 0.79]). The genetic algorithm produced a 6-item subset from the SCL-90-R, which produced comparable classification accuracy to the somatisation scores (AUC = 0.73, 95%CI = [0.64, 0.82]).

**Significance:** Compared to patients with ES, patients with PNES report greater symptoms of somatisation, general anxiety, and phobic anxiety against a background of generally elevated psychopathology. While self-reported psychopathology scores are not accurate enough for diagnosis in isolation, elevated psychopathology in these domains should raise the suspicion of PNES in clinical settings.

## 1. Introduction

Two main categories of seizures are commonly diagnosed: epileptic seizures (ES) and psychogenic non-epileptic seizures (PNES). While both ES and PNES are defined as “paroxysmal, time-limited, alterations in motor, sensory, autonomic, and/or cognitive signs and symptoms”[1], PNES have psychological underpinnings, and are not associated with ictal epileptiform activity on EEG[1]. A diagnosis of PNES is made through the exclusion of epilepsy and other organic causes of seizure. As both PNES and ES are paroxysmal in nature, however, inter-ictal EEG cannot necessarily be used to differentiate them, unless the ictus is captured on EEG[1]. While Video-EEG monitoring (VEM) is the gold standard for PNES diagnosis, it involves significant cost and time commitment[2].

Because diagnosis of PNES is difficult, misdiagnosis or delayed diagnosis of PNES is common, with one study finding an average delay in PNES diagnosis of 7 years[3]. The misdiagnosis itself has the potential to cause significant adverse consequences. The delay in diagnosis can cause a significant emotional toll[4], may cause PNES patients to be incorrectly prescribed anti-epileptic medications (AEDs), and may cause the concept of an organic disease to become entrenched in the patient’s conceptualisation of their illness. Aside from the issues associated with polypharmacy, AEDs are generally ineffective in PNES and may worsen seizures[5]. As such, there is a clear need for accurate and inexpensive screening tools that might assist in identifying patients with PNES, allowing for early consideration of psychiatric management and more targeted and cost-effective VEM use.

One potential patient characteristic that may differentiate patients with PNES and ES is the prevalence of comorbid psychopathology. Multiple studies have suggested increased prevalence of psychiatric comorbidity in ES patients[6–9] compared to the general population, and one meta-analysis has found an increased risk of psychiatric comorbidity in PNES compared to ES[10]. Specifically, the existing literature suggests an increased prevalence of depression[11] and anxiety[6] in ES patients compared to healthy controls. There is a high prevalence of depression[12], anxiety[13], and somatisation[14] in patients with PNES compared to the normal population, as well as evidence for increased levels of depression[12], anxiety[13–17] and somatisation[14] in PNES patients compared to ES patients. Taken together, this suggests that some domains of psychopathology may potentially discriminate between ES and PNES patients. As a result, psychopathology levels, either in general or in association with particular domains, presents a potential differentiating factor between ES and PNES patients.

As psychopathology measurements have the potential to distinguish ES and PNES patients, psychiatric symptomatology questionnaires may have potential utility in screening for PNES. Several studies have suggested that long, multi-domain psychiatric symptomatology questionnaires, such as the Minnesota Multiphasic Personality Inventory (MMPI) and the Personality Assessment Inventory (PAI), may potentially distinguish ES and PNES[18,19]. Previous literature has also examined the profile changes in smaller single-domain questionnaires, such as the Hospital Anxiety and Depression Scale[13]; however, studies using these shorter questionnaires have not explored the classification accuracy of any profile changes, even when statistically significant differences were reported between patient groups. This suggests a potential gap in the available literature, however one potential reason for this may be that a multi-domain psychopathology measurement could provide improved classification accuracy and hold more clinical utility.

Medium-length multi-domain questionnaires are an alternative form of psychopathology measurement, which retains the benefits of measuring multiple psychopathology domains, while also being less time-consuming than longer measurement instruments. One example of a medium-length questionnaire is the Symptom Checklist 90-Revised (SCL-90-R)[20], which contains 90 Likert-scale questions, compared to 567 true-false questions in the second generation MMPI (MMPI-2). Few studies have examined the profile changes in this questionnaire, and no studies have examined the clinical utility of this questionnaire in distinguishing ES and PNES patients. This presents a gap in the literature.

Our study aimed to examine the profile differences between PNES and ES patients on individual SCL-90-R domain scores, as well as a general measure of psychopathology derived from dimensional reduction of the SCL-90-R’s domains. A secondary aim was to determine the diagnostic utility of the SCL-90-R in distinguishing patients with ES and PNES. Finally, we aimed to used machine learning methods, in the form of a genetic algorithm search tool, to find a subset of SCL-90-R questionnaire items that demonstrate the highest diagnostic utility for PNES.

## 2. Methods

### 2.1 Participants

Participants were patients admitted to the VEM unit at the Royal Melbourne Hospital, Australia between 2002 and 2017. Patients were included if they: completed the SCL-90-R during admission as part of routine clinical care; had completed demographics documented in the medical files; had adequate English language ability to complete psychological questionnaires; were aged greater than 13 (as the SCL-90-R is validated for age 13 and over); and did not have a major psychiatric or neurocognitive impairment (e.g., intellectual disability, delirium, severe dyslexia, active psychosis) that would prevent completion of psychological questionnaires. This study was approved by the Melbourne Health Human Ethics Committee (HREC#: QA2012044).

### 2.2 Clinical Diagnosis

VEM consensus diagnosis was made by multidisciplinary team meeting of neurologists, neuropsychiatrists, neuropsychologists, neuroradiologists, and neurophysiology scientists. The patient’s clinical history, VEM results, neurological examination, neuropsychiatric assessment, and neuroimaging findings (where applicable) were all considered when formulating a final diagnosis. Based on the VEM diagnosis, patients were categorised into one of four groups: epilepsy, PNES, mixed (epilepsy + PNES), and non-diagnostic. The non-diagnostic group was comprised of patients who did not have a clear diagnosis from VEM, or experienced events that were neither epileptic seizures nor PNES (eg. cardiovascular events, vasovagal syncope, panic attacks). Patients categorised in the epilepsy or mixed groups were further divided into subgroups based on individual epilepsy syndromes: temporal lobe epilepsy (TLE), extra-temporal focal epilepsy, or generalised epilepsy. Patients in the mixed (epilepsy + PNES) group were excluded from analysis, in order to examine the specific profile differences between epilepsy and PNES. Patients in the non-diagnostic group were also excluded due to their heterogeneous nature, and the inability to classify their events. These patients were included in the sample characteristics to provide a full appreciation of the study population.

### 2.3 Questionnaires

The Symptom Checklist 90 Revised (SCL-90-R) was used to assess individual domains of psychopathology[20]. The SCL-90-R is a 90-item five-point Likert scale self-report psychiatric symptom questionnaire, with ratings provided on 9 primary symptom dimensions. These dimensions were: somatisation, obsessive-compulsive, interpersonal sensitivity, depression, anxiety, hostility, phobic anxiety, paranoid ideation, psychoticism. The internal consistency of the subdomain scores is high (⍰ = 0.77-0.90) and the test-retest reliability is also adequate (0.68-0.83). The validity of the SCL-90-R has been extensively investigated by the test authors[20] and has been shown to be suitable for clinical use.

For each domain of psychopathology as listed in the SCL-90-R, individual domain Z-scores were calculated for each patient, using SCL-90-R nonpatient mean and standard deviation groups, as published in the test manual[20]. These scores provided a representation of the degree of symptomatology in each patient relative to the normal population. SCL-90-R scores were not known to the clinical team at the time of VEM diagnosis, and therefore did not inform the consensus diagnosis.

### 2.4 Statistical Analysis

Statistical analysis was performed using the *R* and *Jamovi* software packages[21,22].

#### 2.4.1 Linear Models

First, Bayesian general linear mixed models (GLMMs) were computed to investigate the differences in psychopathology profiles between patients with epilepsy and patients with PNES. A model selection approach was used to determine whether specific terms should be included in the model. Age and gender were included in all models as covariates. Univariate models were specified as:

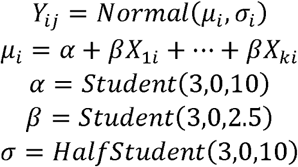

where *Y*_ij_ is the outcome variable *j* for person *i*. Multilevel models included a random intercept for participant (distinguished by patient UR). Model parameters were estimated using a full Bayesian approach in the *brms* package for R[23,24]. Hamiltonian MCMC was implemented using the *stan* sampler[25]. Point estimates were extracted as the median of the posterior distribution for each predictor, with 95% highest density intervals (HDIs). A Bayesian equivalent of the coefficient of determination (*R*^2^) was computed[26]. Care was taken to inspect the posterior predictive distributions to evaluate model fit. The posterior predictive probability value (PPP) was computed for each model. Values between 0.05 and 0.95 were considered reasonable[27]. Effect sizes, in the form of Cohen’s *d*[28] were computed where relevant. All main analyses were also replicated using frequentist methods (not shown).

#### 2.4.2 Principal Components Analysis

Next, principal components analysis (PCA) was performed to determine the presence of a single psychopathology factor *p*, which provided a maximal explanation of variance. The number of components was determined objectively using parallel analysis. This approach compares the observed eigenvalues to the eigenvalues obtained by performing PCA on a matrix of random values of the same dimensionality as the input matrix. Component scores were saved from the resulting components and used in subsequent analyses. Bayesian GLMs were then used, as described above, to determine the differences in *p* between patients with epilepsy and patients with PNES.

#### 2.4.3 Determining Classification Accuracy

Receiver operating characteristic curves (ROC curves) were computed in order to investigate classification accuracy of the SCL-90-R in detecting PNES patients. The *pROC* package[29] was used for all analyses. The optimal cut-off score for each variables was computed using Youden’s method[30]. Cut-off scores were also computed that produced a sensitivity closest to 90% in order to evaluate the SCL-90-R from a screening perspective. For each threshold, sensitivity, specificity, PPV and NPV were calculated, as well as their associated 95% confidence intervals using bootstrapping.

#### 2.4.4 Determining an Optimal Subset of SCL-90-R Items

In order to determine whether a specific subset of SCL-90-R items produced greater clinical utility in detecting PNES, a genetic algorithm (GA) search tool[31] was used to determine which combination of individual symptom items produced the greatest classification accuracy. The dataset of PNES-only and ES-only patients was randomly split into a 70% training sample and a 30% testing sample. The area under the curve (AUC) was used as the cost function. A population size of 300 was used for each generation, with a maximum of 1000 generations per run. The simulation was allowed to run for a maximum of 50 generations without the best fitness level changing before it was terminated. This procedure was repeated 100 times and items that were selected >50% of the time were selected as final subset of items. Total scores were then computed for these items and analysed using a Bayesian GLM as described above. This was then repeated in the testing data set.

## 3. Results

### 3.1 Sample Characteristics

In total, 628 patients met inclusion criteria and were included in the study. Of these patients, 293 (47% of total) were diagnosed with ES only, 158 (25%) with PNES only, 31 (5%) with both ES and PNES, and 146 (23%) were considered non-diagnostic. Within the epilepsy group, the majority of patients had a diagnosis of Temporal Lobe Epilepsy (TLE) (*n =* 157, 54%), followed by Extra-Temporal Focal Epilepsy (*n =* 79, 27%), and Generalised Epilepsy (*n =* 66, 23%). It is also important to note that due to gaps in availability of clinical data, 1 patient (<1%) did not have a specified epilepsy type, and 12 patients (<1%) did not have a recorded focal epilepsy type.

As shown in *Table 1*, the mean age in the PNES group was lower than all other groups. Gender also differed between diagnostic groups, with the PNES group having a higher proportion of females than all other groups. Only participants from the PNES and ES groups were retained for subsequent analyses.

**Table 1.** Sample characteristics by diagnosis, including demographic information and SCL-90-R domain Z-scores.

|  | *Mean (SD)* |  |  |  |
| --- | --- | --- | --- | --- |
|  | *ES*<br>(*n* = 293) | *PNES*<br>(*n* = 158) | *Mixed*<br>(*n* = 31) | *Non-Diagnostic*<br>(*n* = 146) |
| *Demographics* |  |  |  |  |
| Age | 38.74 (14.94) | 34.02 (13.16) | 39.88 (12.79) | 40.69 (16.61) |
| Female (n (%)) | 169 (57.7%) | 119 (75.3%) | 23 (74.1%) | 90 (61.6%) |
| *SCL-90-R domains* |  |  |  |  |
| Anxiety (ANX) | 1.22 (2.03) | 2.48 (2.61) | 2.66 (2.42) | 1.22 (2.18) |
| Depression (DEP) | 1.47 (1.84) | 2.51 (2.27) | 2.75 (2.13) | 1.39 (2.10) |
| Hostility (HOS) | 0.87 (1.92) | 1.51 (2.23) | 1.62 (2.08) | 0.84 (1.95) |
| Interpersonal Sensitivity (IS) | 1.29 (1.98) | 2.13 (2.52) | 2.48 (2.36) | 1.24 (2.20) |
| Obsessive-Compulsive (OCD) | 1.93 (2.00) | 2.64 (2.15) | 2.84 (2.08) | 1.76 (2.14) |
| Paranoia (PAR) | 0.59 (1.62) | 1.39 (2.10) | 1.67 (2.11) | 0.58 (1.72) |
| Phobic Anxiety (PHO) | 1.30 (2.72) | 3.42 (3.80) | 3.14 (2.70) | 1.37 (2.58) |
| Psychoticism (PSY) | 1.65 (2.55) | 2.44 (2.96) | 3.19 (2.99) | 1.36 (2.42) |
| Somatisation (SOM) | 1.03 (1.67) | 2.67 (2.96) | 1.83 (1.63) | 1.56 (1.90) |

### 3.2 Difference in SCL-90-R profiles between PNES and ES

Bayesian linear mixed models (GLMMs) were computed to investigate whether domain scores on the SCL-90-R differed between groups. The best fitting model included age, gender, and the diagnostic group by SCL-90-R domain interaction (*Table S1*). As shown in *Table 2*, there was evidence for a main effect of age, with overall domain scores declining over the life-span (*b* = -0.02, 95%HDI = [-0.03, -0.01]). There was also a main effect of diagnostic group, indicating that patients in the PNES group had higher overall domain scores compared to patients in the ES group (*b* = 0.75, 95%HDI = [0.28, 1.18]). There was no evidence for a main effect for gender (*b* = 0.24, 95%HDI = [-0.15, -0.59]).

**Table 2:** Bayesian general linear mixed model results

| *Model term* | *Main effect*<br>*b [95% HDI]* | *Interaction effect*<br>*b [95% HDI]* |
| --- | --- | --- |
| *Demographics* |  |  |
| Age | -0.02 [-0.03, -0.01] | - |
| Gender (Male) | 0.24 [-0.15, 0.59] | - |
| PNES | 0.75 [0.28, 1.18] | - |
| *SCL-90-R domains* |  |  |
| Anxiety (ANX) | -0.43 [-0.65, -0.22] | 0.47 [0.10, 0.80] |
| Depression (DEP) | -0.18 [-0.39, 0.03] | 0.24 [-0.1, 0.61] |
| Hostility (HOS) | -0.78 [-0.99, -0.57] | -0.15 [-0.51, 0.2] |
| Interpersonal Sensitivity (IS) | -0.36 [-0.57, -0.15] | 0.05 [-0.32, 0.40] |
| Obsessive-Compulsive (OCD) | 0.28 [0.06, 0.48] | -0.08 [-0.45, 0.26] |
| Paranoia (PAR) | -1.06 [-1.27, -0.84] | 0.00 [-0.36, 0.35] |
| Phobic Anxiety (PHO) | -0.35 [-0.56, -0.13] | 1.32 [0.98, 1.69] |
| Somatisation (SOM) | -0.61 [-0.83, -0.40] | 0.84 [0.49, 1.20] |

**Note:** The psychoticism domain (PSY) was used as the reference class for contrast coding.

There was evidence for an interaction effect between diagnostic group and domain scores on three domains: anxiety (*b* = 0.47, 95%HDI = [0.10, 0.80]), *d* = 1.22 [large]), phobic anxiety (*b* = 1.32, 95%HDI = [0.98, 1.69], *d* = 2.08 [large]) and somatisation (*b* = 0.84, 95%HDI = [0.49, 1.20], *d* = 1.59 [large]). The mean scores for each group by domain are shown in *Figure 1*, while *Figure 2* shows the between-group effect sizes.

**Figure 1:**
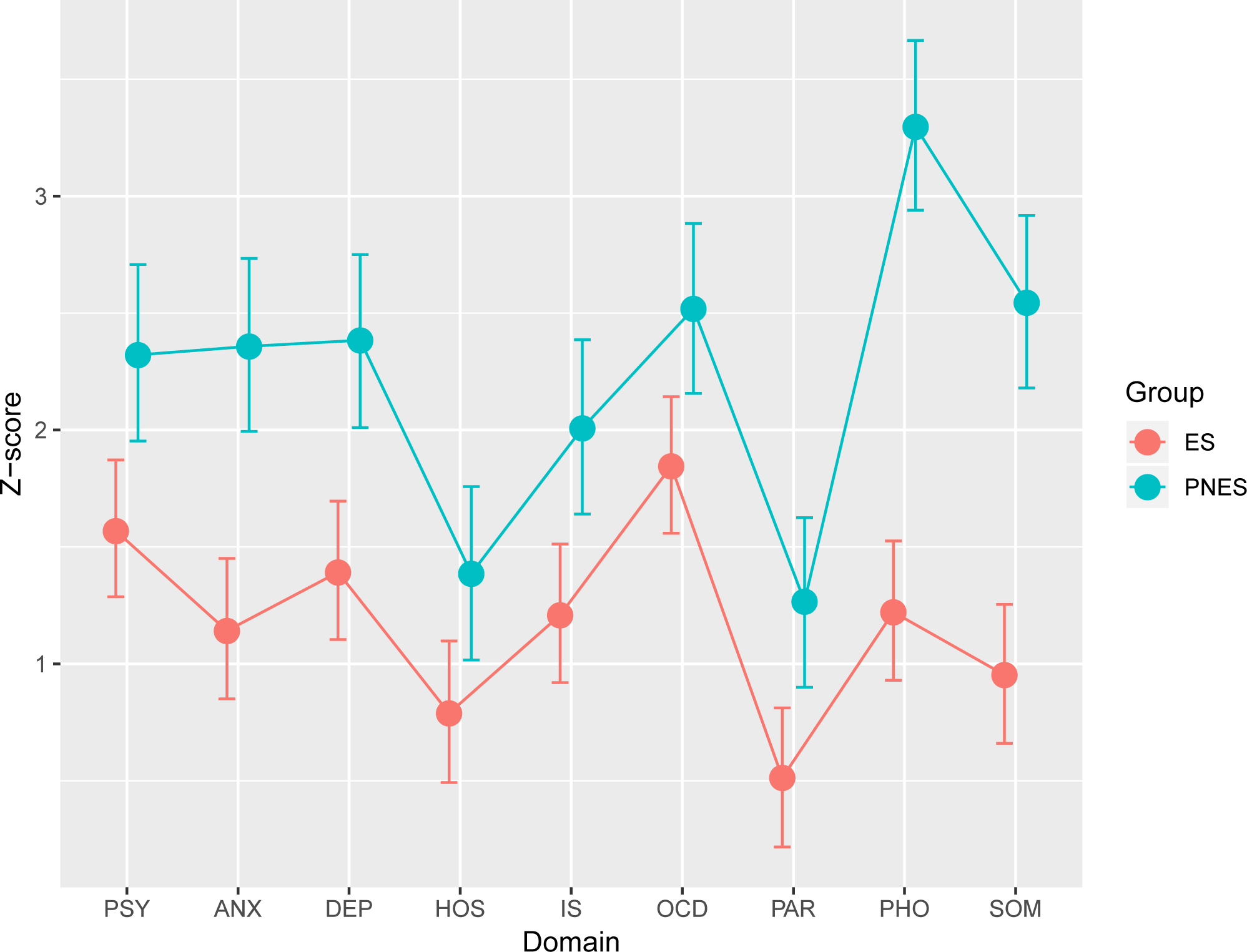
Bayesian marginal effects plots. Marginal effects plots of individual domain scores by patient group (ES, PNES). As shown, patients with PNES were elevated in all domains, with specific elevations in general anxiety, phobic anxiety, and somatisation. PSY: psychoticism, ANX: anxiety, DEP: depression, HOS: hostility, IS: interpersonal sensitivity, OCD: obsessive-compulsive, PAR: paranoia, PHO: phobic anxiety, SOM: somatisation.

**Figure 2:**
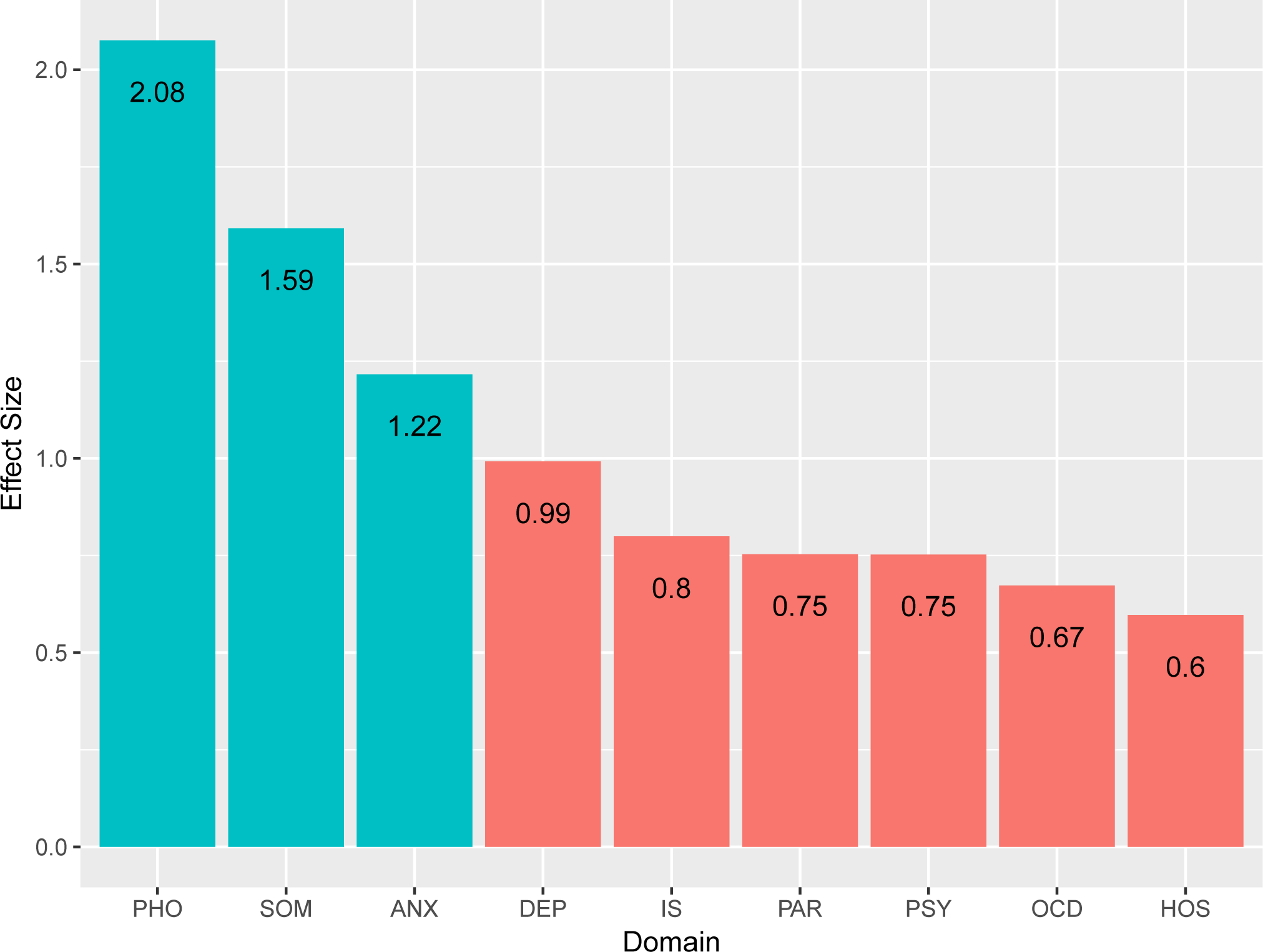
Standardised effect sizes for each domain comparing PNES and ES patients. PSY: psychoticism, ANX: anxiety, DEP: depression, HOS: hostility, IS: interpersonal sensitivity, OCD: obsessive-compulsive, PAR: paranoia, PHO: phobic anxiety, SOM: somatisation.

### 3.3 Common Psychopathology Factor p

Principal components analysis demonstrated the presence of a single *p* component that explained 74% of the total variance of the individual domain scores. A Bayesian GLM revealed a main effect of diagnostic group (*b* = 1.35, 95%HDI = [0.86, 1.82]), with PNES patients having higher scores compared to ES patients (*d* = 1.35 [large]). There is evidence for a negative relationship with age (*b* = -0.03, 95%HDI = [-0.04, -0.01]), but no evidence for an effect of gender (*b* = 0.33, 95%HDI = [-0.13, 0.82]).

### 3.4 Classification Accuracy of Domain Scores and p

Receiver operating characteristic (ROC) curves were computed to investigate the diagnostic accuracy of each SCL-90-R domain score and the *p* factor (*Figure 3A*). As shown in *Table 3*, all areas under the curve (AUCs) were statistically significant. The somatisation domain produced the best classification (AUC = 0.74, 95%CI = [0.69, 0.79]), with an optimal sensitivity of 0.65 (95%CI = [0.54, 0.78]) and a specificity of 0.76 (95%CI = [0.63, 0.85]). The generalised psychopathology factor *p* also produced better-than-chance classification accuracy (AUC = 0.67, 95%CI = [0.61, 0.72]), with sensitivity of 0.67 (95%CI = [0.45, 0.80]) and a specificity of 0.76 (95%CI = [0.47, 0.82]).

**Table 3:**
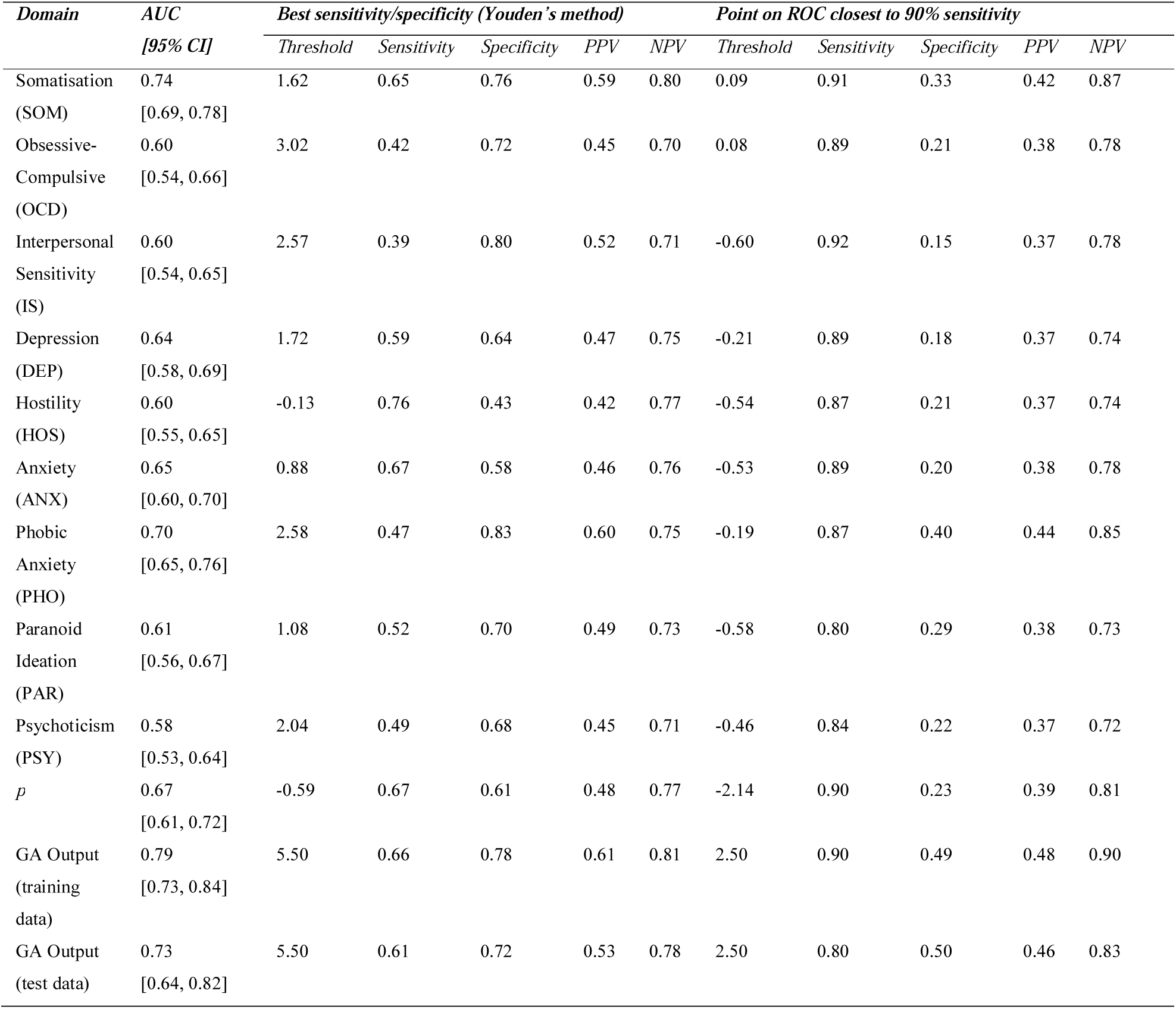
ROC Curve Statistics

| *Domain* | *AUC* | *Best sensitivity/specificity (Youden's method)* |  |  |  |  | *Point on ROC closest to 90% sensitivity* |  |  |  |  |
| --- | --- | --- | --- | --- | --- | --- | --- | --- | --- | --- | --- |
|  | *[95% CI]* | *Threshold* | *Sensitivity* | *Specificity* | *PPV* | *NPV* | *Threshold* | *Sensitivity* | *Specificity* | *PPV* | *NPV* |
| Somatisation (SOM) | 0.74<br>[0.69, 0.78] | 1.62 | 0.65 | 0.76 | 0.59 | 0.80 | 0.09 | 0.91 | 0.33 | 0.42 | 0.87 |
| Obsessive-Compulsive (OCD) | 0.60<br>[0.54, 0.66] | 3.02 | 0.42 | 0.72 | 0.45 | 0.70 | 0.08 | 0.89 | 0.21 | 0.38 | 0.78 |
| Interpersonal Sensitivity (IS) | 0.60<br>[0.54, 0.65] | 2.57 | 0.39 | 0.80 | 0.52 | 0.71 | -0.60 | 0.92 | 0.15 | 0.37 | 0.78 |
| Depression (DEP) | 0.64<br>[0.58, 0.69] | 1.72 | 0.59 | 0.64 | 0.47 | 0.75 | -0.21 | 0.89 | 0.18 | 0.37 | 0.74 |
| Hostility (HOS) | 0.60<br>[0.55, 0.65] | -0.13 | 0.76 | 0.43 | 0.42 | 0.77 | -0.54 | 0.87 | 0.21 | 0.37 | 0.74 |
| Anxiety (ANX) | 0.65<br>[0.60, 0.70] | 0.88 | 0.67 | 0.58 | 0.46 | 0.76 | -0.53 | 0.89 | 0.20 | 0.38 | 0.78 |
| Phobic Anxiety (PHO) | 0.70<br>[0.65, 0.76] | 2.58 | 0.47 | 0.83 | 0.60 | 0.75 | -0.19 | 0.87 | 0.40 | 0.44 | 0.85 |
| Paranoid Ideation (PAR) | 0.61<br>[0.56, 0.67] | 1.08 | 0.52 | 0.70 | 0.49 | 0.73 | -0.58 | 0.80 | 0.29 | 0.38 | 0.73 |
| Psychoticism (PSY) | 0.58<br>[0.53, 0.64] | 2.04 | 0.49 | 0.68 | 0.45 | 0.71 | -0.46 | 0.84 | 0.22 | 0.37 | 0.72 |
| *p* | 0.67<br>[0.61, 0.72] | -0.59 | 0.67 | 0.61 | 0.48 | 0.77 | -2.14 | 0.90 | 0.23 | 0.39 | 0.81 |
| GA Output (training data) | 0.79<br>[0.73, 0.84] | 5.50 | 0.66 | 0.78 | 0.61 | 0.81 | 2.50 | 0.90 | 0.49 | 0.48 | 0.90 |
| GA Output (test data) | 0.73<br>[0.64, 0.82] | 5.50 | 0.61 | 0.72 | 0.53 | 0.78 | 2.50 | 0.80 | 0.50 | 0.46 | 0.83 |

**Figure 3:**
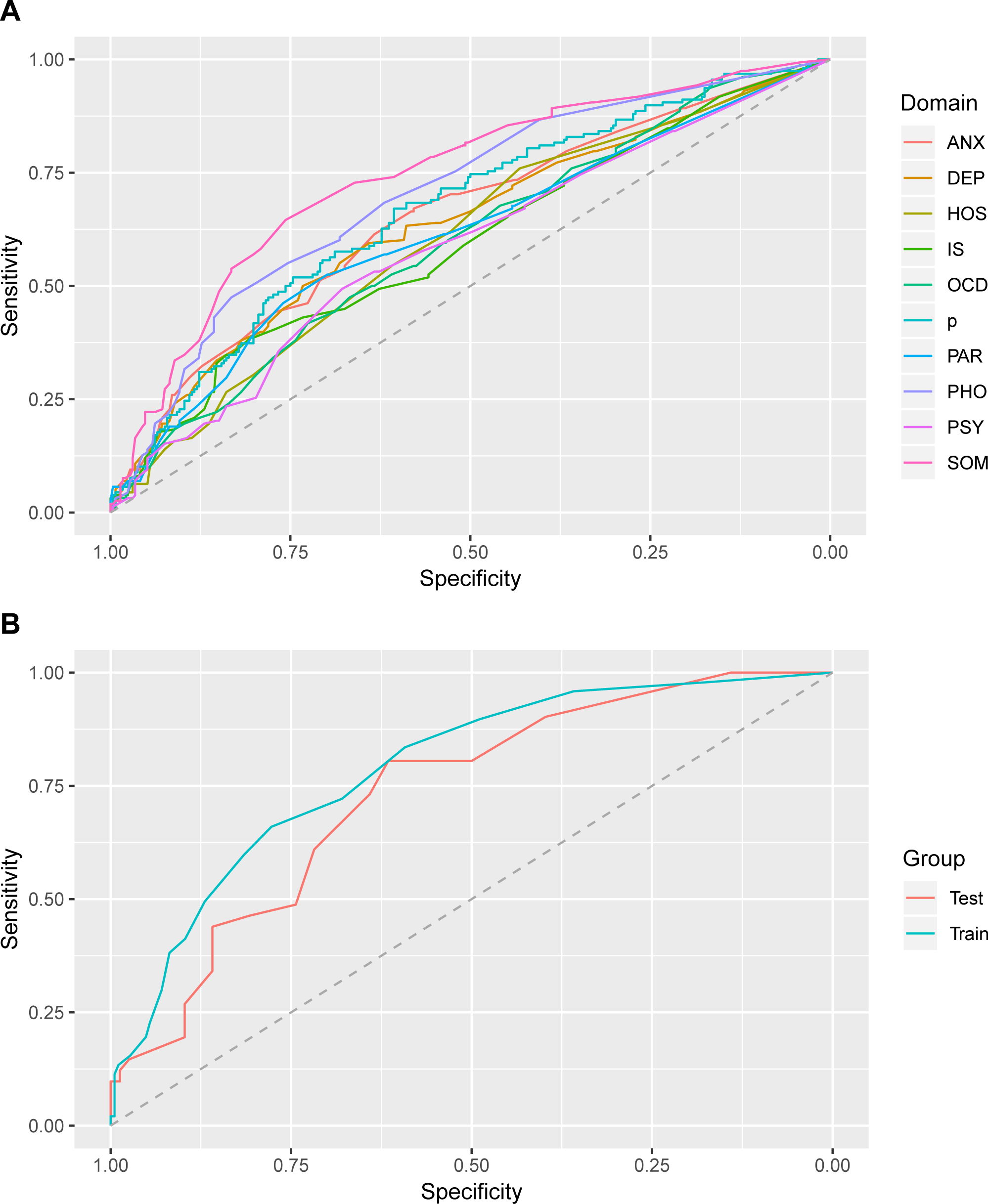
Receiver operator characteristic (ROC) curves for individual domain scores and the general psychopathology factor p (A), as well as the genetic algorithm output scores (B), including both the 70% training dataset and the 30% testing (cross-validation) dataset. PSY: psychoticism, ANX: anxiety, DEP: depression, HOS: hostility, IS: interpersonal sensitivity, OCD: obsessive-compulsive, PAR: paranoia, PHO: phobic anxiety, SOM: somatisation.

### 3.5 Determining an Optimal SCL-90-R item subset for Detecting PNES

The genetic algorithm produced a set of 6 SCL-90-R symptom items which were present in the output of greater than 50 of the 100 runs. These included items from the domains of generalised anxiety, phobic anxiety and somatisation (question numbers 1, 4, 12, 25, 70, 86). McDonald’s omega was 0.86, suggesting that the items chosen had high psychometric reliability.

For each patient, a genetic algorithm output score was produced by taking the sum of the numeric responses from each of the 6 SCL-90-R symptom items. Receiver operating characteristic (ROC) curves were computed to investigate the diagnostic accuracy of this output score in both the test and training subsets of the patient cohort (*Figure 3B*). As shown in Table 3 this score significantly discriminated between PNES and ES cases, and outperformed the somatisation domain score in the training set (AUC = 0.79, 95%CI = [0.73, 0.84]), with a sensitivity of 0.66 (95%CI = [0.58, 0.91]) and a specificity of 0.78 (95%CI = [0.52, 0.85]). The score remained a significant classifier in the test set (AUC = 0.73, 95%CI = [0.64, 0.82]) with a sensitivity of 0.61 (95%CI = [0.46, 0.76]) and a specificity of 0.72 (95%CI = [0.62, 0.82]). Classification accuracy of the test set was similar to the somatisation domain score.

## 4. Discussion, Conclusions

In this study we examined the multivariate psychopathology profiles of a large group of patients with either PNES or ES. A number of key findings emerged. First, we found evidence for a general psychopathology factor (otherwise known as *p*) which explained the majority of variance in psychopathology symptoms. This general psychopathology factor was elevated in patients with PNES compared to those with ES, which suggests an elevated general propensity for psychological distress in patients with PNES. Second, we found that certain domains of psychopathology (in particular, the domains of general anxiety, phobic anxiety, and somatisation) were elevated in PNES even when taking into account increased levels of general psychological distress. All associated effect sizes were large, suggesting potential clinical utility in identifying patients at risk of being diagnosed with PNES.

The finding of a single psychopathology factor is novel in epilepsy research, and the elevation of this *p* factor in patient with PNES compared to ES is consistent with previous research conducted in this population. Specifically, several studies have reported increased levels of generalised psychological distress in PNES patients[32–34], and a recent meta-analysis revealed high rates of psychiatric comorbidity in this patient group[10]. Taken together, our finding of an elevated *p* factor confirms a general propensity for elevated psychological distress in patients with PNES.

The finding of elevated anxiety, phobic anxiety and somatisation symptoms in PNES patients is also consistent with previous literature, both when examining psychopathology incidence and response profiles on other psychopathology symptom questionnaires. Previous work has suggested an increased prevalence of anxiety disorders in PNES patients compared to ES patients[13–17], as well as a particular increased prevalence of post-traumatic stress disorder[15,35]. While some studies did not find a significant change in anxiety disorder prevalence in the PNES population[36,37], the majority of studies were limited by smaller sample sizes. The association between PNES and higher levels of somatisation has also been extensively explored. Several studies have reported increased somatoform disorder prevalence in PNES patients[14,36] as well as elevated levels of somatisation symptomatology[18,19,38–43]. It is important to note that PNES is itself categorised in the DSM-V as a subtype of somatic symptom disorder[44]. In this context, our finding of elevated general somatisation syndromes in PNES suggests similarities between PNES and other functional neurological disorders.

One apparent conflict between our results and previous research is the lack of evidence for elevation of certain symptom domains in PNES relative to ES. For example, we did not find a specific elevation in depressive symptomatology in the PNES group. A previous systematic review found a significant increase in depression prevalence in PNES patients, compared to both ES and control patients[12], and previous studies using psychopathology questionnaires suggested elevated depressive symptomatology in PNES patients[13,16,19,32,40,43,45]. An explanation for this apparent conflict is that we controlled for the *general* propensity towards elevated psychopathology when examining differences on individual domains. This was achieved by including a main effect of diagnostic group in our linear models and then examining the interaction terms between psychopathology domain and diagnostic group. These results suggest that there is overall greater psychopathology in patients with PNES, which was confirmed by our discovery of a common *p* factor. Many previous studies have investigated psychopathology symptoms in isolation, without considering a global increase in the propensity towards psychopathology in PNES. Our findings suggest that, in the presence of an elevated general psychopathology factor, any single psychopathology domain considered in isolation will appear elevated. Only the domains of generalised anxiety, phobic anxiety and somatisation were found to be elevated against this general increase in psychopathology.

A third key finding from our study related to the PNES classification accuracy of both individual psychopathology domain scores, as well as the calculated general psychopathology factor *p*. While many studies have reported group differences between PNES and ES in terms of psychopathology, few have specifically examined the ability for such differences to translate into screening or diagnostic strategies. This is essential if psychopathology questionnaires are to be incorporated into routine clinical workflows. Our findings suggest that while PNES and ES patients have significantly different psychopathology profiles, the classification accuracy of individual domain scores and the general psychopathology factor *p* is not sufficiently accurate for use as a diagnostic test. Our finding of low classification performance is consistent with previous literature in this area. While some studies have claimed that some clinical scales in various questionnaires display increased PNES classification accuracy[19,46], upon cross-validation in other studies the majority of these measures have not shown sufficient accuracy for use in clinical diagnosis[47–49]. One potential reason for this is that the PNES population demonstrates heterogeneity in terms of symptom, risk factor and comorbidity profiles, suggesting that one questionnaire response profile may not completely describe all patients with PNES. One study showed evidence for this by demonstrating the presence of different ‘clusters’ of PNES questionnaire response profiles[50]. A second reason for insufficient accuracy may be the particular questionnaire used in this study. Multiple studies have used different questionnaires, which may each capture their own pertinent aspects of PNES psychopathology. One study found that PNES classification accuracy was improved when combining elements from multiple different psychopathology questionnaires (in this case, the Minnesota Multiphasic Personality Inventory and the Personality Assessment Inventory), rather than using a single questionnaire alone[39]. A third reason for lack of diagnostic accuracy may involve the limitations of using self-report instruments alone. While self-report instruments have been shown to demonstrate good accuracy in major depressive disorder symptomatology assessment [51], using multiple assessment methods (including structured clinical interviews) may reduce the risk of under- or over-predicting levels of psychopathology [52].Such an approach, however, would be time consuming and might not be appropriate for use in screening applications.

A final finding of our study involved the use of a genetic algorithm to narrow down our set of predictors to those which had the highest PNES classification accuracy. The 6 items returned from the algorithm were from the anxiety, phobic anxiety, and somatisation dimensions, concurring with the evidence provided by the linear models of changes in these domain scores between PNES and ES patients. However, while these items were shown to have slightly improved classification accuracy on the training set on which it was built, cross-validation with the testing dataset found classification accuracy to be slightly poorer than that of the somatisation domain alone. As such, even when using advanced machine learning approaches to classification, it was not possible to derive a highly accurate screening or diagnostic approach appropriate for clinical translation.

Overall, our study demonstrated a range of strengths. A large sample was obtained, all of whom had been diagnosed using gold-standard VEM as the majority of other methods have not proven reliable for PNES classification[1]. Unlike previous studies utilising the SCL-90-R[32–34], both general psychopathology factors and individual psychopathology domains were examined, and the general psychopathology factor was calculated through dimensional reduction. Other methods, including a genetic algorithm, were also utilised to maximise the potential classification accuracy. It should be noted that determining classification accuracy itself is often an overlooked aspect when examining group differences in clinical research.

However, it is also apparent that some limitations are present in the study. As this study was conducted retrospectively, some clinical variables other than those previously described were not available or not reported in the majority of patient data. It is important to note that some questionnaire subscales demonstrated improved classification accuracy when combined with other data[38,41,53], such as EEG measurements and clinical variables (including seizure length and length of time since first seizure). As such, provision of a large study combining both questionnaire results and other variables is a potential direction for future research. In addition, although we excluded patients who did not have sufficient cognitive performance to complete the SCL-90-R, we did not have sufficient data to investigate the effect of cognitive performance on psychopathology. This study also used a patient cohort from a single tertiary centre, which poses the risk of the results not being generalisable if this cohort is not representative of the general population.

In conclusion, our study demonstrated robust and large differences between patients with ES and PNES using a comprehensive and validated psychopathology instrument. There was a generally increased propensity for psychopathology in patients with PNES, as well as specific elevations in general anxiety, phobic anxiety, and somatisation. While these differences were not large enough to sufficiently classify patients, they do suggest that elevated psychopathology, particularly in the somatisation domain, should raise the suspicion for PNES in patients undergoing VEM. These results point towards some psychopathology domains (notably those of general anxiety, phobic anxiety, and somatisation) as potential avenues of research for future psychometric detection methods, as well as the potential for higher classification accuracy when using questionnaire results in combination with other clinical variables.

## Data Availability

Data available at https://doi.org/10.17605/OSF.IO/GQRYT.

## 6. List of Abbreviations

ES: epileptic seizures
PNES: psychogenic non-epileptic seizures
TLE: temporal lobe epilepsy
VEM: video-EEG monitoring
EEG: electroencephalogram
AED: anti-epileptic medication / anti-epileptic drug
HDI: highest density interval
AUC: area under the curve (used in ROC Curves)
CI: confidence interval
GLMM: general linear mixed models
GLM: general linear model
PCA: principal components analysis
ROC Curves: receiver operating characteristic curves
GA: genetic algorithm
SCL-90-R: Symptom Checklist 90-Revised

## SCL-90-R Domain Abbreviations

PSY: psychoticism
ANX: anxiety
DEP: depression
HOS: hostility
IS: interpersonal sensitivity
OCD: obsessive-compulsive
PAR: paranoia
PHO: phobic anxiety
SOM: somatisation
MMPI: Minnesota Multiphasic Personality Inventory
PAI: Personality Assessment Inventory
DSM-V: Diagnostic and Statistical Manual of Mental Disorders, Fifth Edition

## 6. Acknowledgements

We would like to thank all patients admitted to the Royal Melbourne Hospital’s VEM unit who have taken the time to participate in this study. We also would like to extend our appreciation to all staff who have assisted in the collection of data.

